# Improving the Diagnosis of Small Intestinal Bacterial Overgrowth Using an At-Home Handheld App Connected Breath Analysis Device (AIRE)

**DOI:** 10.1101/2022.04.21.22274143

**Authors:** Guillermo Barahona, Barry Mc Bride, Áine Moran, Sahar Hawamdeh, Luisa Villatoro, Robert Burns, Bo Konings, Robert Bulat, Megan McKnight, Claire Shortt, Pankaj J. Pasricha

## Abstract

**INTRODUCTION:** Small Intestinal Bacterial Overgrowth (SIBO) is a common yet underdiagnosed condition. Lactulose hydrogen breath tests (LHBT) are typically used to detect SIBO; however, current breath testing methods require specialised, expensive equipment and technical support and are either done at a point-of-care facility and/or have to be mailed to a central laboratory. To address these issues a novel hand-held breath analyzer (AIRE®, FoodMarble) was tested. The aims of this study were first, to perform a technical assessment of the AIRE device, second to compare the performance of the AIRE device against a commercially available mail-in LHBT kit using a zero-inflated negative binomial mixed effect model.

**METHODS:** Three AIRE devices were tested with certified test gases covering a diagnostically meaningful range (hydrogen mixed with air at 3 ppm, 10 ppm and 50 ppm). For the clinical study, 36 patients suspected to have SIBO presenting to a tertiary level clinic were provided with an AIRE device and performed concurrent LHBTs at home with a mail-in LHBT kit.

**RESULTS:** The overall average readings (mean ± SD) for the AIRE devices tested at 3 ppm, 10 ppm and 50 ppm H_2_ were: 3.5 ± 0.7 ppm; 10.7 ± 1.1 ppm and 49.5 ± 2.6 ppm respectively. The overall mean absolute error across the tested devices was 1.2 ppm. A significant positive correlation (r = 0.78, p < 0.001) was demonstrated between AIRE and mail-in kit H_2_ values.

**DISCUSSION:** The AIRE device is a compelling alternative to mail-in LHBT kits for the diagnosis of SIBO. The AIRE device may also offer advantages over other traditional breath testing methods.

## INTRODUCTION

Small intestinal bacterial overgrowth (SIBO) is defined as a condition in which an abnormally high amount of coliform bacteria are present in the small bowel. This overgrowth can result in premature fermentation of carbohydrates before reaching the colon (1). Commonly recognized causes include gastric achlorhydria (i.e. due to longstanding proton pump inhibitor (PPI) use), post-surgical bowel stasis, and gastrointestinal motility disorders leading to bowel stasis. SIBO is associated with a range of non-specific gastrointestinal (GI) symptoms. These include bloating, abdominal pain, distension and diarrhoea. If left undiagnosed, it can lead to weight loss and nutritional deficiencies (2).

SIBO overlaps with several GI conditions, however its true prevalence is unknown. It can be diagnosed using small bowel aspirate culture showing 10^3^ – 10^5^ colony-forming units (CFU) per mL of duodenal aspirate (3). However, this method is invasive, costly, subject to contamination and the diagnostic criterion varies by orders of magnitude. Breath testing is a simpler, more cost effective, non-invasive diagnostic method. Hydrogen breath testing relies on the demonstration of an early rise in breath hydrogen in response to an orally ingested carbohydrate, typically lactulose or glucose (4).

A lactulose hydrogen breath test (LHBT) is typically carried out using either a hospital based bench-top breath analyser or a mail-in LHBT kit. Hospital based breath testing requires specialized, expensive equipment and technical staff to carry out testing with each patient, which can take up to 5 hours. Mail-in, LHBT kits require patients to breathe into, seal and manually label a series of sample bags or tubes. Samples are then posted to a laboratory for analysis. Extended time between sample collection and analysis can result in sample leakage, due to the diffusion of the hydrogen through the material of the collection system and the increased humidity of the breath sample (5). Therefore, it is preferable to collect and analyse breath samples in real-time.

To address these issues, we tested an app-connected, novel hand-held breath analyzer (AIRE®, FoodMarble) that allows patients to perform H_2_ breath testing and simultaneous symptom monitoring from the home. The AIRE device has previously been validated in two studies comparing it to the gold standard hospital-based breath test device (6, 7). The aims of this study were first, to perform a technical assessment of the AIRE device, second to compare the performance of the AIRE device against a commercially available mail-in LHBT kit using simple linear regression and a zero-inflated negative binomial mixed effect model.

## METHODS

### Technical Assessment

The technical performance and repeatability of the AIRE device was determined by an ISO 17025:2017 accredited testing and calibration laboratory (GAS Analysis Services, Wicklow, Ireland). Certified test gases (hydrogen mixed with air at 3 ppm, 10 ppm and 50 ppm) were analysed using a gas chromatography machine to verify their concentrations. The test gases were heated and humidified to replicate human breath (24°C ≤ temperature ≤ 28°C, 80% ≤ relative humidity ≤ 95%). The test gases were passed through each AIRE device for five seconds to match real world operation. Each device was tested three times using each test gas sequentially, with a 15 minute gap between each test to replicate a typical LHBT testing scenario. Three AIRE devices were tested three times at each test gas concentration on three non-consecutive days.

### Study Subjects

The study population consisted of adult patients experiencing chronic (3 months or more) gastrointestinal symptoms including nausea, bloating, distention, altered bowel movements, weight loss or abdominal pain with no evidence of mechanical obstruction. Subjects were required to have received a clinical diagnosis of SIBO with intent to obtain a LHBT. Exclusion criteria included a history of inflammatory bowel disease or antibiotic use within 30 days prior to study commencement. The study was registered with ClinicalTrials.gov (NCT04309396).

### Performance of LHBT

Subjects performed simultaneous LHBTs at-home using the AIRE device and a commercially available mail-in kit. LHBTs were performed in a standardised manner using the instructions provided by the manufacturer. In line with established clinical guidelines a positive LHBT was defined as a ≥ 20 ppm rise in breath H_2_ above baseline within 90 minutes (8). Subjects were required to follow a diet low in fermentable carbohydrates for the 24 hours before the test. They underwent a minimum of 12 hours fasting, where only water was permitted. On the morning of the LHBT, subjects checked their fasting breath H_2_ levels using the AIRE device and app. If their fasting breath H_2_ was elevated (> 15 ppm), they were advised to re-check it in 20 minutes, for up to one hour. If their fasting breath H_2_ remained elevated, they were advised to postpone the test until the following morning. A baseline breath reading was recorded on the AIRE device and using the mail-in kit. The test substrate, 15 mL of lactulose, was ingested. Every 15 minutes a breath test was recorded on the AIRE device and the mail-in kit for a total of two hours. Upon completion of the LHBT, the AIRE results were immediately available for review by a gastroenterologist using the FoodMarble online dashboard. The commercial test kit was mailed by the patient to a testing lab for the measurement of the breath test samples. Results were received by PDF within 3 – 10 days.

### Statistical Analysis

Correlations between breath H_2_ concentrations from the AIRE devices and mail-in kits were determined using Pearson’s correlation and simple linear regression analysis. This analysis was performed using IBM SPSS software version 28.

In order to model test kit performance, we constructed a zero-inflated negative binomial mixed-effect model, implying independence between H_2_ ppm readings when conditioned on all fixed and random effects. In particular, in addition to an “always zero” mixture component, the model was specified with a second mixture component specifying method (i.e. AIRE or mail-in kit), time and an intercept term as fixed effects as well as random effects for method, time and the intercept for each subject in the study such that:

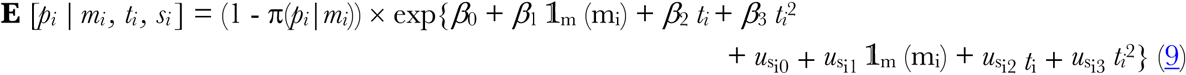

where

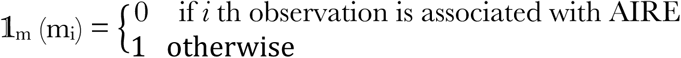

and

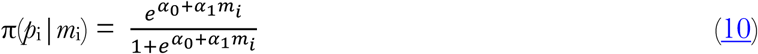

Where *p*_i_, *m*_i_, *t*_*i*_, and *s*_i_ correspond to H_2_ ppm reading, method, time and subject respectively for observation *i*; π(*p*_i_|*m*_i_) corresponds to the probability that *p*_*i*_ comes from the “always zero” mixture component. *β* _j_ and 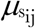 correspond to fixed effects and subject-wise random effects respectively for subject *i* and for *j* ∈ 1,…,3. The model was fitted using a glmmTMB software package (9) and DHARMa software (10) was used to assess the resulting model fit.

## RESULTS

### Technical Assessment

Three AIRE devices were tested with three certified tests gases covering a diagnostically significant range of H_2_ concentrations (3 ppm, 10 ppm and 50 ppm hydrogen mixed with air). The overall average readings for the three AIRE devices tested at 3 ppm, 10 ppm and 50 ppm hydrogen were as follows: 3.5 ± 0.7 ppm (mean ± SD); 10.7 ± 1.1 ppm (mean ± SD) and 49.5 ± 2.6 ppm (mean ± SD) respectively. To test day-to-day repeatability, the protocol was repeated on three non-consecutive days (Figure 1). Across all three AIRE devices the mean absolute error (MAE) at 3 ppm, 10 ppm and 50 ppm were: 0.7 ppm, 1 ppm and 1.9 ppm respectively. The overall MAE was 1.2 ppm.

**Figure 1.**
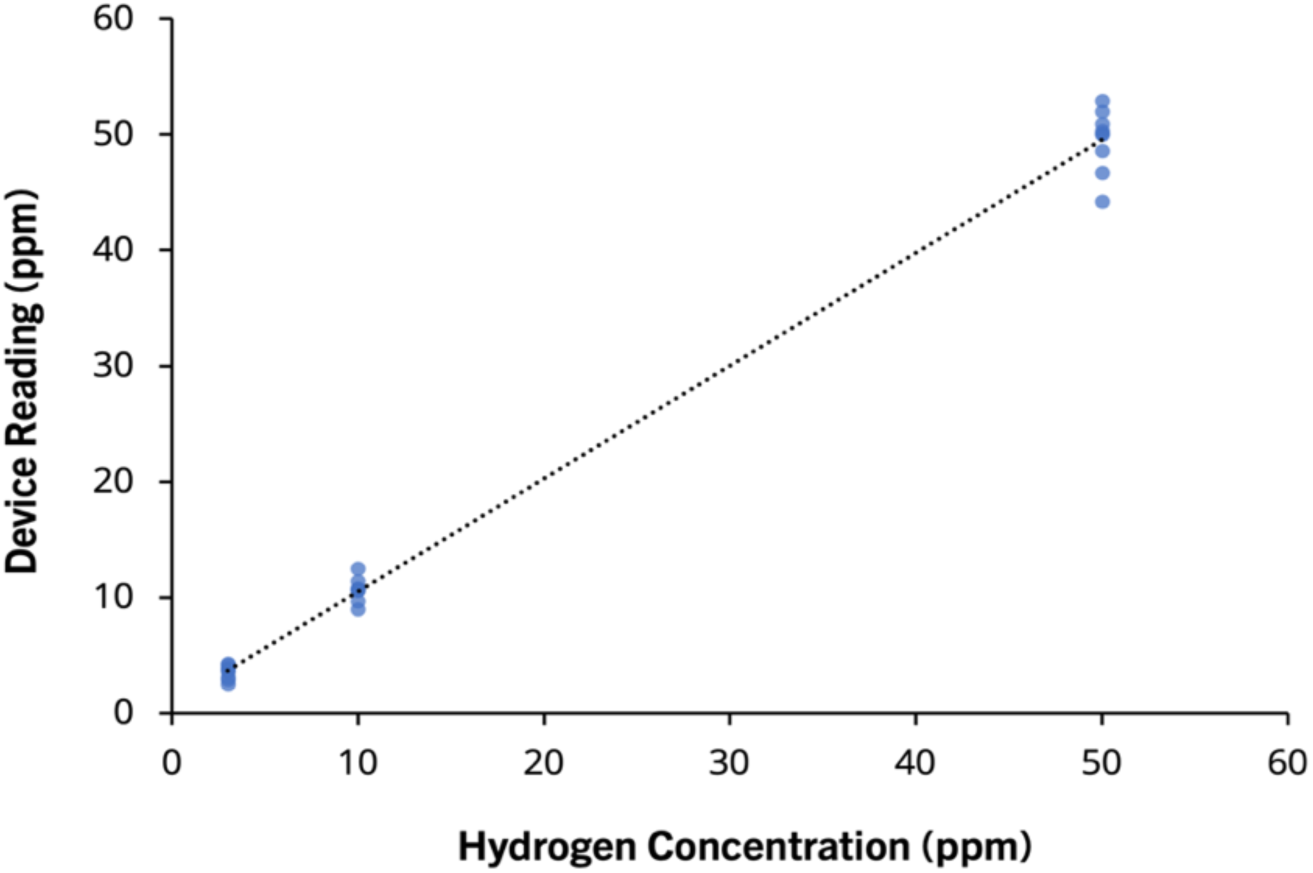
Linearity and repeatability curve for three AIRE devices tested at 3 ppm, 10 ppm and 50 ppm H_2_/air on three non-consecutive days.

**Figure 2.**
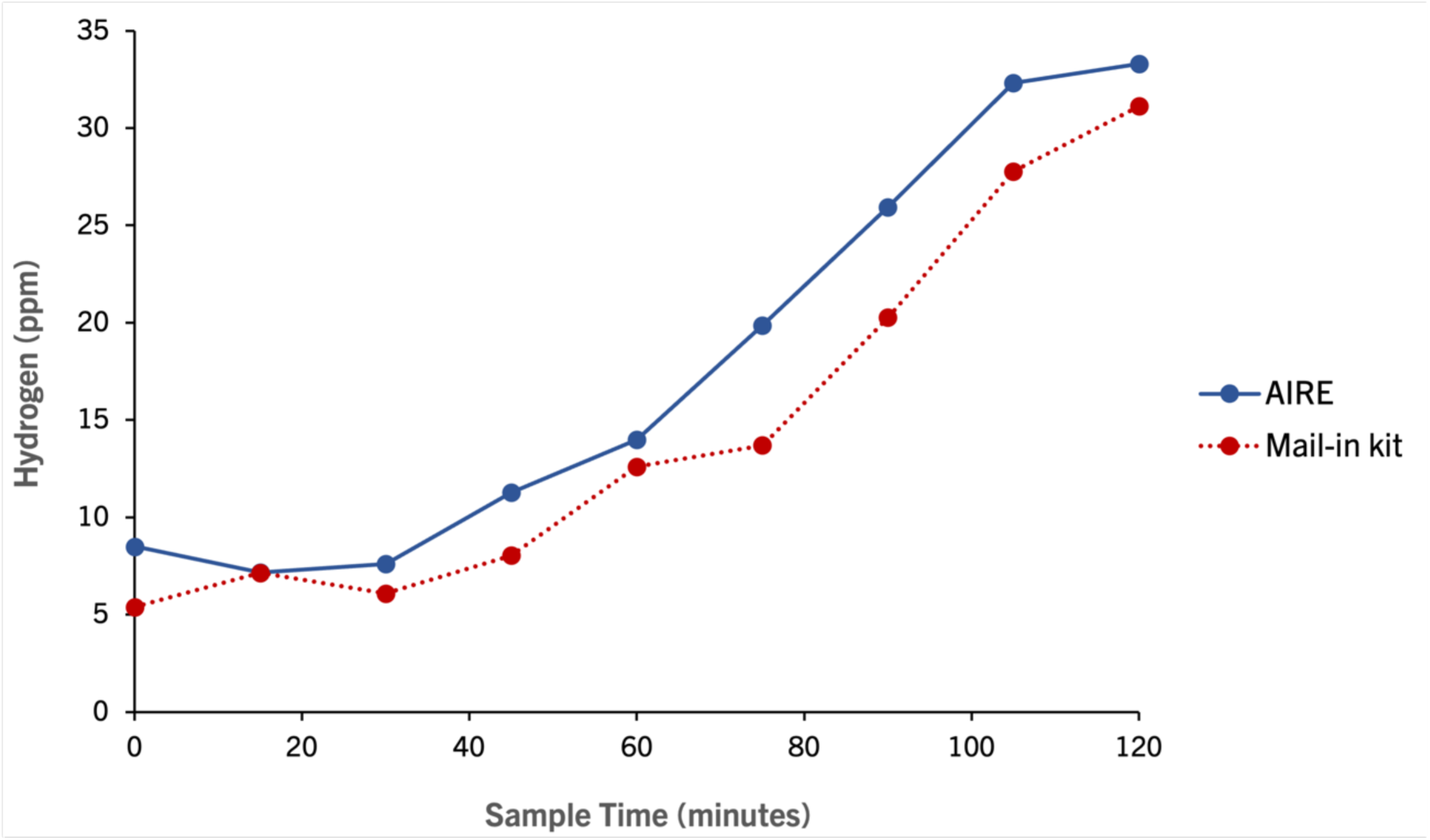
Mean breath H_2_ concentrations collected during the LHBT using AIRE and the mail-in kit (n=36).

### Comparison of LHBT performance using AIRE and mail-in kit

All subjects performed LHBTs using the standard diagnostic method. 33% (12/36) of LHBTs performed with the AIRE device were positive. 25% (9/36) of LHBTs performed with the mail-in kit were positive (Table 1). There was diagnostic agreement in 86% (31/36) of cases.

**Table 1.** LHBT comparison of AIRE and mail-in kit (n=36). LHBT positive ≥ 20 ppm rise in H2. LHBT negative ≤ 20 ppm rise in H_2_.

|  | LHBT Positive | LHBT Negative |
| --- | --- | --- |
| AIRE | 12 | 24 |
| Mail-in kit | 9 | 27 |

Of the five cases which were in diagnostic disagreement, four were positive on the AIRE device and negative with the mail-in kit while one was positive with the mail-in kit and negative on the AIRE device. Of the four LHBTs that were positive on the AIRE device and negative with the mail-in kit, two showed a rise in breath H_2_ ≥ 20 ppm within 90 minutes on the AIRE device while the H_2_ readings with the mail-in kit flatlined.

The average (mean ± SD) baseline breath H_2_ values measured with the AIRE device and mail-in kit were 8.5 ± 7.4 ppm and 5.4 ± 6.5 ppm respectively. The average (mean ± SD) peak H_2_ values measured were 39.2 ± 32.8 ppm and 33.9 ± 34.6 ppm on the AIRE device and mail-in kit respectively. The overall mean breath H_2_ values measured were 18.6 ± 24.9 ppm and 15.2 ± 23.3 ppm on the AIRE device and mail-in kit respectively (Table 2).

**Table 2.** Mean baseline, overall and peak breath H_2_ ppm for AIRE devices and mail-in kits (n=36).

|  | AIRE | Mail-in kit |
| --- | --- | --- |
| Baseline $H_2$ ppm (mean ± SD) | 8.5 ± 7.4 | 5.4 ± 6.5 |
| Overall $H_2$ ppm (mean ± SD) | 18.6 ± 24.9 | 15.2 ± 23.3 |
| Peak $H_2$ ppm (mean ± SD) | 39.2 ± 32.8 | 33.9 ± 34.6 |

A total of 293 pairs of breath measurements taken during 36 LHBTs were included in the linear regression analysis. The results demonstrated a significant positive linear correlation between the breath H_2_ measurements using the AIRE device and the mail-in kits (r = 0.78, p < 0.001) (Table 3).

**Table 3.**
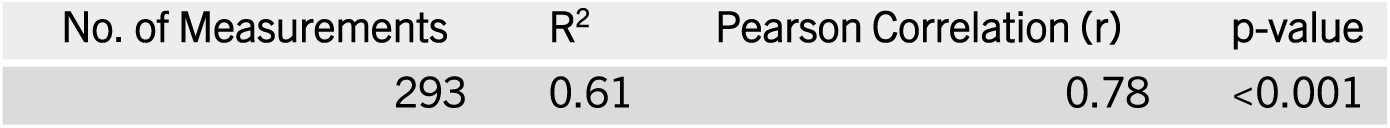
Linear regression analysis for the association of breath H_2_ measurement using AIRE and mail-in kits (n=36).

The zero-inflated negative binomial mixed-effect linear model suggested that, on average, mail-in kits produced lower H_2_ ppm readings than the AIRE devices (Wald z-test p-value: 0.012; log-mean estimate: -0.59; standard error: 0.23; 95% confidence interval: -1.05 — -0.13). On average, mail-in kits were also more likely to produce zero readings (Wald z-test p-value: 0.017; log-odds estimate: 2.74; standard error: 1.15; 95% confidence interval: 0.49 — 4.99).

## DISCUSSION

Digital health technologies have made the remote monitoring of health metrics increasingly possible. Breath testing is commonly used in the diagnosis and management of functional digestive disorders. However, traditional breath testing requires the patient to travel to the clinic in a fasting state, and only gathers a single snapshot view of what is happening. The provision of a personal breath testing device to each patient would enable them to perform repeat tests from the home, which has the potential to improve treatment and management of the patient. Accordingly, in the present study the performance of an app-connected personal breath tester (AIRE®, FoodMarble) was compared to that of a commercially available mail-in kit.

There was a high degree of diagnostic agreement (86%) between both breath measurement methods. The overall positive yield for the LHBTs performed using AIRE (33%) was comparable to that reported elsewhere for both lactulose and glucose breath testing in similar patient cohorts (11, 12). There was also a significant positive linear correlation between both methods (r=0.78, p < 0.001). These data support the use of the AIRE device as a viable alternative to mail-in LHBT kits for the “standard” diagnosis of SIBO based on breath H_2_ concentrations in response to lactulose.

Of the four LHBTs that were positive on the AIRE device and negative with the mail-in kit, two had peak hydrogen readings of 26 ppm and 29 ppm above baseline within 90 minutes during the LHBT performed on the AIRE device. The corresponding mail-in kits for these LHBTs reported flatline H_2_ readings. Furthermore, 6% (19/324) of all breath tests collected using the mail-in kits were reported as invalid or insignificant samples. This was due to sample bags having insufficient volumes of sample gas for analysis. Previously, *Beauchamp et al*. demonstrated that the levels of some volatile organic compounds collected in gas sampling bags, like H_2_ gas, decreased exponentially over time (5). These losses were thought to be due to diffusion through the outer material of the collection bags resulting in samples with unacceptable percentage recoveries after 70 hours of storage. The samples from the mail-in test kit, that were deemed invalid, or those that flatlined may have been due to sample bag leakage. This hypothesis is supported by the zero-inflated negative binomial mixed-effect model, which suggested that the mail-in kits produced lower H_2_ readings and were more likely to produce zero readings on average.

The leakage effect is generally more pronounced in samples with high humidity (5). In the current study, time between collection and reporting of results for the mail-in kits ranged from 3 to 10 days. Given the diffuse nature of H_2_ and the high humidity of breath samples, this may make leakage more likely to occur. Furthermore, the Rome Consensus Conference working group have recommended that once breath samples have been taken they should be analysed within 6 hours (4). The ability to analyse breath readings in real time would mitigate this risk.

The technical assessment of the AIRE device using calibrated test gases resulted in an overall MAE of 1.2 ppm across the three tested devices, with good day-to-day repeatability. These results indicate that the AIRE device has the technical capability to perform multiple LHBTs over time. The substrates used for breath testing have varying degrees of suitability depending on a patients motility, with glucose being more suited to those with faster motility (1, 13, 14). Therefore, the capacity for the patient to perform a repeat, confirmatory test using an AIRE device, using an alternate substrate (lactulose or glucose) may be advantageous.

The present study was intended to prove the equivalence of a simple and patient-friendly device to a commercial mail-in test in the diagnosis of SIBO and we believe this primary objective was achieved. Additionally, in some instances, AIRE provided greater levels of accuracy due to the real-time measurement of samples. Furthermore, an independent, technical validation of AIRE demonstrated that it could accurately and repeatably measure known, certified gas concentrations over a diagnostically meaningful range (3 ppm, 10 ppm and 50 ppm). These data further support the use of the AIRE device for multiple, repeat LHBT tests. Performing multiple LHBT’s, either prior to treatment or post treatment is not typically done due to limited access to breath testing. However providing a patient with their own device, would enable them to test as often as needed, in a more cost effective, patient friendly manner. Borderline LHBT results are common and so, retesting may provide more confidence in a diagnosis and may result in a reduction in antibiotic misuse. Furthermore, in patients whose symptoms do not fully resolve after antibiotic use or recur after a period of time, repeating the test also provides useful information either on the persistence of SIBO if positive or eliminating it as a cause of symptoms, if negative (15, 16).

Notable benefits of the AIRE device include: a clinician dashboard that displays results in real-time; the patient is able to check their fasting breath measurement prior to commencing the test (if baseline breath H_2_ levels are above 15 ppm, patients are asked not to proceed with the LHBT, and to recheck every 20 minutes, up to one hour); test protocol compliance (breath reminder notifications are sent to the patient every 15 minutes for the test duration) and automatic symptom tracking after each breath test. Post-test symptom tracking is also possible. The AIRE device therefore provides an alternative way to measure exhaled H_2_ that has the potential to address many of the limitations of standard breath testing.

## Data Availability

All data produced in the present study are available upon reasonable request to the authors.

